# The influence of sex steroid treatment on insular connectivity in gender dysphoria

**DOI:** 10.1101/2022.01.21.22269471

**Authors:** Murray B Reed, Patricia A Handschuh, Manfred Klöbl, Melisande E Konadu, Ulrike Kaufmann, Andreas Hahn, Georg S Kranz, Marie Spies, Rupert Lanzenberger

**Author notes:** Correspondence to: Prof. Rupert Lanzenberger, MD, PD, Medical University of Vienna, Department of Psychiatry and Psychotherapy, Austria. contributed equally (Murray B Reed, Patricia A Handschuh).

## Abstract

The influence of sex hormones on brain function has been investigated in multiple neuroimaging studies. Sexually dimorphic characteristics were found for the insular cortex, though little is known about hormonal effects on sex-specific functional connectivity patterns and insular functions ranging from emotion regulation to interoception and higher-level cognition. Thus, better understanding of direct sex steroid effects on insular connectivity remains essential. Thereby, gender-dysphoric individuals receiving gender-affirming hormone therapy represent an interesting cohort to address this gap in available knowledge.

To analyze the potential effect of sex steroids on insular connectivity at rest, 14 transgender women, 19 transgender men, 24 cisgender women, and 15 cisgender men were recruited. All participants underwent two magnetic resonance imaging sessions involving resting-state acquisitions separated by a median time period of 4.5 months. Between scans, transgender subjects received gender-affirming hormone therapy.

A seed based functional connectivity analysis revealed a significant 2-way interaction effect of group-by-time between right insula, cingulum, left middle frontal gyrus and left angular gyrus. Post-hoc tests revealed an increase in connectivity for transgender women when compared to cisgender men. Furthermore, spectral dynamic causal modelling showed reduced effective connectivity from the posterior cingulum and left angular gyrus to the left middle frontal gyrus as well as from the right insula to the left middle frontal gyrus.

These findings suggest a considerable influence of long-term estrogen administration and androgen suppression on brain networks implicated in interoception, own-body perception and higher-level cognition. Nevertheless, further studies are needed to shed light on the underlying mechanisms.

## 1. Introduction

The insula is located within the lateral sulcus of both hemispheres and covered by the frontoparietal and the temporal opercula. Given its afferent and efferent connections to multiple brain regions (e.g., the thalamus (1, 2), limbic (3-5) and cortical regions (6, 7)), it is associated with a wide range of functional processes.

The insula’s broad structural connectivity is reflective of its wide range of functions. Highly dynamic processes involving the interaction of emotion and cognition (e.g., decision making) have been linked to the insula, encompassing the regulation of explicit emotional experiences and the integration of somatosensory as well as autonomic stimuli in the context of cognitive processing (8, 9). Studies using functional magnetic resonance imaging (fMRI) demonstrate functional and structural differences between insular subdivisions (10). As summarized by Uddin et al., meta-analyses on coactivation patterns of the insular cortex suggest the primary involvement of the ventral anterior part in affective regulation, whereas the dorsal anterior part is rather implicated in cognitive processing (11). Sensorimotor activation and interoceptive functions are largely attributed to the posterior insula (12-14). However, this tripartite parcellation is called into question by functional heterogeneity within certain subdivisions as well as the dense interconnection across them (10).

Alterations of the insula could be linked to a wide range of neuropsychiatric conditions. In a meta-analysis of voxel-based morphometry including 193 studies comprising scans from over 15800 patients suffering from schizophrenia, bipolar disorder, depression, addiction, obsessive-compulsive disorder or anxiety, Goodkind and colleagues found that the dorsal anterior cingulate, right insula, and left insula showed reductions of gray matter volume in all diagnostic categories, accompanied by poor executive functioning when compared to healthy controls. (15). Furthermore, the insula is suggested to be involved in the pathomechanism of affective disorders (16-18), probably due to its connection to regions implicated in emotion regulation (19). Additionally, pathologic alterations of the insula were found in substance use disorder (20), schizophrenia (21), autism spectrum disorder (22, 23) as well as in the context of non-motor and neuropsychiatric symptoms of Parkinson’s disease and various forms of dementia (24).

Looking at the brain as an organ with sexually dimorphic characteristics at every age, the influence of sex hormones (25-27) and non-hormonal effects of genes found on the X and Y chromosomes (28) have been addressed in various neuroimaging studies, underlining typical divergences between the brain of men and women, both in structure and function. Such differences were also found regarding the insula. Next to distinctive structural characteristics (29), several gender-related functional dissimilarities could be shown, e.g., in language (30) and affective processing (31), potentially playing a role for a wide range of neuropsychiatric conditions. In a structural MRI study, Klabunde and colleagues reported gender-related differences of the anterior circular sulcus of the insula in boys suffering from PTSD symptoms, showing larger volume and surface area than their healthy male counterparts. In contrast, smaller average volume and surface area could be detected in girls with PTSD when compared to control girls (32). Other morbidities associated with gender-specific insular differences are major depressive disorder (33), chronic pain (34), substance abuse (35) or migraine (36), just to name a few.

A unique possibility to further investigate the effects of sex hormones on brain structure and connectivity is given by cohorts diagnosed with gender dysphoria (GD) undergoing gender-affirming hormone therapy (GHT). As concisely reviewed by Kranz et al., GHT was shown to modulate brain structure and function in numerous ways (37). However, little is known about the effect of GHT on the insula, despite previous reports on its sex-specific characteristics. On a molecular level, Kranz et al. found a statistically significant reduction of insular MAO-A distribution volume in transgender men receiving exogenous testosterone (38), probably contributing to functional changes driven by altered neurotransmitter metabolism. A few fMRI studies reporting on sex-specific differences of insular connectivity were published, though sample sizes were small and highly varying study designs were used (39, 40).

The present study was implemented to investigate the potential influence of long-term GHT administration on insular connectivity at rest across four study groups encompassing transgender women and transgender men as well as two control groups comprising cisgender women and cisgender men.

## 2. Methods and Materials

This study was conducted according to the Declaration of Helsinki including all current revisions and the good scientific practice guidelines of the Medical University of Vienna. The protocol was approved by the institutional review board (EK Nr.: 1104/2016) and registered at clinicaltrials.gov (NCT02715232).

### 2.1. Study Design

The study was conducted in a longitudinal design. All participants underwent two MRI sessions separated by a median of 4.5 months. Each session included structural, diffusion-weighted, task-based and resting-state MRI as well as spectroscopy. During the time between the two MRI sessions, transgender subjects received an individualized GHT treatment regimen that comprised the administration of testosterone and progestins (if menstruation still occurred) in transgender male participants, whereas transgender women received a steroidal antiandrogen and estradiol. Hormone levels were determined by blood draw at each MRI session.

#### Gender-affirming hormone therapy

Hormone therapy was conducted according to the following protocol implemented at the Department of Obstetrics & Gynecology, Division of Gynecologic Endocrinology and Reproductive Medicine, Medical University of Vienna, Austria: Transgender men received either 1000mg testosterone undecanoat every 8-16 weeks (Nebido 1000mg/4ml i.m.), or alternatively up to 50mg testosterone daily (Testogel 50mg/day or Testavan 23mg/pump 1-2 pumps/day or testosterone crème as a magistral formula 12,5mg/pump 3-4 pumps/day transdermally). If menstruation still occured, transgender men were treated with either lynestrenol (Orgametril 2-3 tablets/day) or desogestrel daily (Moniq Gynial or Cerazette 0.075mg 1 tablet/day). Transgender women received cyproterone acetate daily (Androcur 50mg/day) and either estradiol transdermally (Estramon 100μg transdermal patch 2x/week or Estrogel gel 0.75-1.5 mg/day) or oral estradiol (Estrofem 2×2mg/day p.o.). Additionally, 2.5 mg of an alpha-5-reductase-inhibitor was administered every second day (Finasterid Actavis/Arcana/Aurobindo) in case of extensive hair loss. In selected cases, subjects received GnRH-analogues like triptorelin (decapeptyl 0.1mg/day subcutaneously).

#### Sex hormones

For monitoring sex hormone levels, luteinizing hormone, follicle-stimulating hormone, progesterone, estradiol, testosterone, sex hormone binding globulin and dehydroepiandrosterone sulfate were determined from the blood drawn before each MRI session. Testosterone, progesterone, and estradiol were used as covariates in the analyses.

### 2.2. Participants

Transgender (TX) individuals seeking GHT were recruited from the Unit for Gender Identity Disorder at the General Hospital in Vienna. Control subjects were recruited via social media, designated message boards at the Medical University of Vienna and at other universities in Vienna. Cisgender men and cisgender women were age and education level matched to transgender men and transgender women, respectively. Inclusion criteria comprised a diagnosis of gender dysphoria according to the Diagnostic and Statistical Manual for Mental Disorders (transgender participants only), version 5 (DSM-5: 302.85) or the International Classification of Diseases, version 10 (ICD-10: F64.1); general health based on medical history, physical examination, electrocardiogram, laboratory screening and structural clinical interview (SCID) for DSM-IV Axis I. All participants gave written informed consent to partake in this study, were insured and received reimbursement for their participation. Participants were excluded in case of major neurological or internal illnesses, pregnancy, any kind of psychiatric diagnosis (cisgender controls) or severe DSM-IV Axis-I comorbidities (transgender subjects), steroid hormone treatment within 6 months prior to inclusion, treatment with psychotropic agents, clinically relevant abnormal laboratory values, MRI contraindications, current substance abuse (excluding smoking), current or past substance-related disorder or non-compliance.

### 2.3. Data acquisition

All MRI data were recorded on a Siemens Prisma 3T scanner using a 64-channel head coil. A whole-brain T1-weighted scan TE / TR = 2.91 / 2000 ms; inversion time = 900 ms; flip angle = 9°; matrix = 240 × 256, 176 slices; 1.0 mm^3^; acquisition time = 7:59 min.

The resting-state data was acquired using the following parameters, TE / TR = 30 / 2050 ms; GRAPPA 2; 210 × 210 mm field of view, 100 × 100 pixel in-plane resolution; 35 axial slices of 2.8 mm (25% gap); flip angle 90°; orientation parallel to the anterior-posterior commissure line.

### 2.4. FMRI preprocessing

Each participant’s data was preprocessed using the following steps: First, physiological artifacts were reduced using PESTICA (41). Further preprocessing steps were performed using SPM12 (https://www.fil.ion.ucl.ac.uk/spm/software/spm12/) using standard parameters. Slice-timing correction was performed to the temporal middle slice, followed by a two-pass realignment of both measurements per subject to the mean image. The images were then normalized to the Montreal Neurological Institute (MNI) standard space and resliced to 2.5 mm^3^. The BrainWavelet Toolbox (42) was used to reduce nonlinear artifact where “chsearch” was set to “harsh” for an increased artifact recognition and “threshold” was set to “20”. The images were gray-matter masked with a custom template and finally smoothed using a Gaussian kernel of three times the resliced voxel size. Further processing steps included a Friston-24 model of motion correction (43) and an adapted compCor approach using an automatically derived number of combined white matter and cerebrospinal fluid regressors (44, 45).

### 2.5. Structural data preprocessing

In parallel, anatomical scans of all participants were preprocessed using the longitudinal recon-all stream (46) and segmented in subject space using FreeSurfer 6. Finally the right insula of each subject was identified. Furthermore, the time courses were then extracted from the each individuals preprocessed RS scans in subject space, corrected for all confounders (despiking, bandpass filter: 0.01-0.1 Hz, CompCor) and finally summarized to its first eigenvariate to match the preprocessing pipeline of the RS data.

### 2.6. Seed based connectivity and dynamic casual modelling

Seed based connectivity (SBC) was calculated using the time course previously extracted from the RS data. Here the anatomically defined right insula was used as the seed and was correlated with each of the RS pipeline processed voxels’ time course. The resulting individual correlation maps were z-transformed and entered into a repeated-measures ANOVA model using SPM12. Interactions of group (FtM, MtF, FC, MC) by time (before and after min. 4 months of hormone therapy) were examined and the results were familywise-error-corrected to p_corr_ ≤ 0.025 (47). The interaction effects were estimated and post-hoc comparisons adjusted again using the Sidak method.

Effective connectivity was assessed using spectral dynamic casual modelling (DCM) implemented in SPM12, where the first temporal eigenvariates of clusters from the SBC analyses surviving multiplicity correction were extracted from the data preprocessed for the DCM analysis (48). Next, fully connected linear spectral two-state DCMs were estimated. The parametric empirical Bayes (PEB) framework in SPM12 was used for group inference. A flat PEB model was employed using free energy. Bayesian model averaging (49) was utilized to prune connections with high uncertainty posterior probability (Pp) < 0.99. Progesterone, testosterone and estradiol were used as covariates in both the repeated-measures ANOVA model and DCM, to account for the individualized GHT. For both the SBC and DCM analyses, each covariate was standardized, and group mean centered before being added to the above-mentioned models.

## 3. Results

Demographics and hormone levels are provided in Table 1. Fewer MtF and matched MC were recruited when compared to FtM and FC. At the second time point, MtF had a lower testosterone and higher progesterone and estradiol levels, whereas FtM had higher testosterone, lower progesterone and lower estradiol levels when compared to MC and FC groups.

**Table 1:**
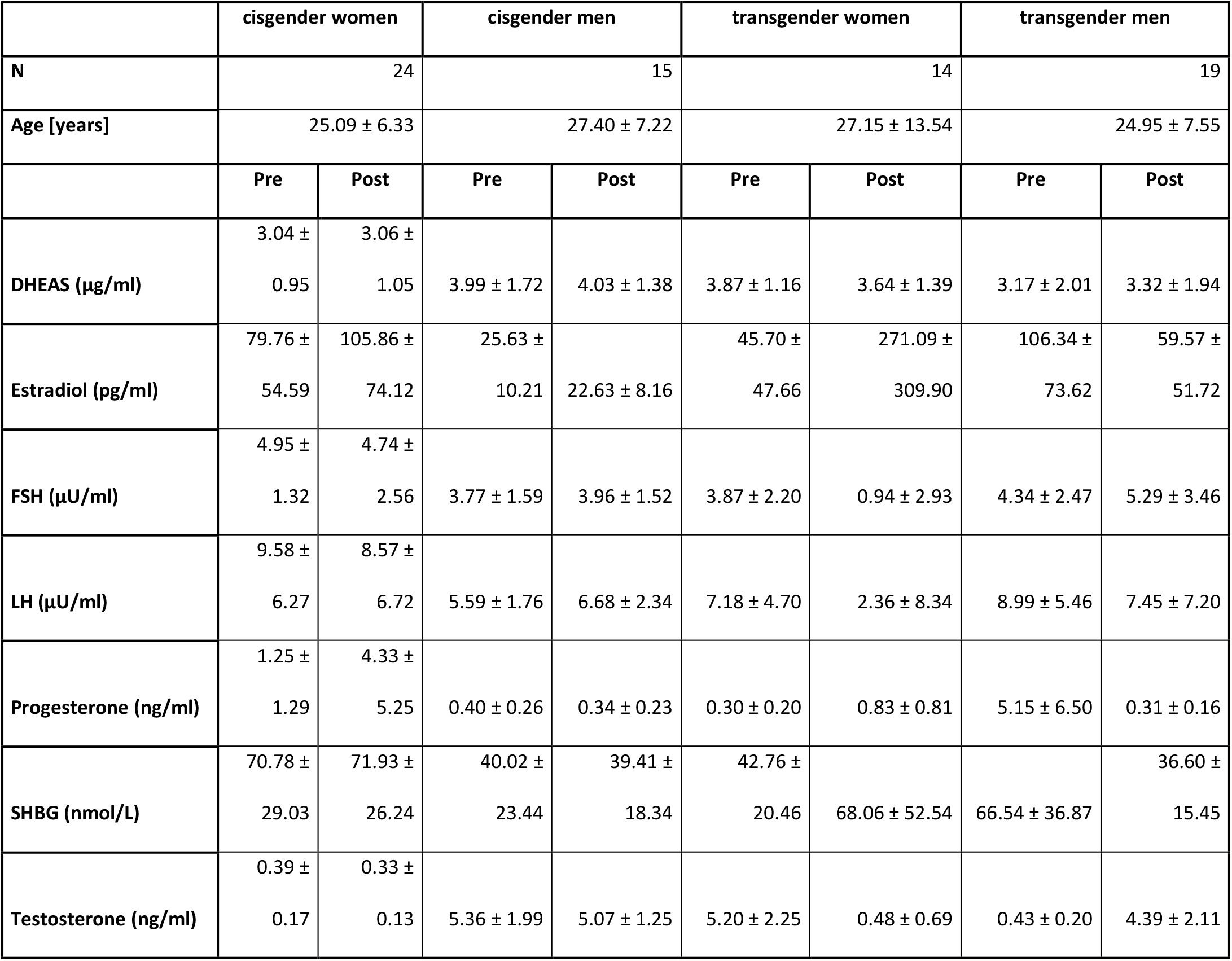
Demographics of all participants and divided into their subgroups. Age is given from the 1st measurement. Hormone levels for the pre-treatment assessment and cisgender participants are provided for comparison only and were not used in the statistical analyses.

### 3.1. Effective and seed-based network connectivity effects

A significant 2-way interaction of group-by-time was discovered. Post-hoc test revealed that the right insula (seed region), cingulum (p_corr_ = 0.047), left middle frontal gyrus (MFG; p_corr_ = 0.037) and left angular gyrus (AG; p_corr_ = 0.028) indicated a greater connectivity in the MtF group when compared to MC (Fig 1a). No significant interaction regarding FtM and FC were found.

**Figure 1:**
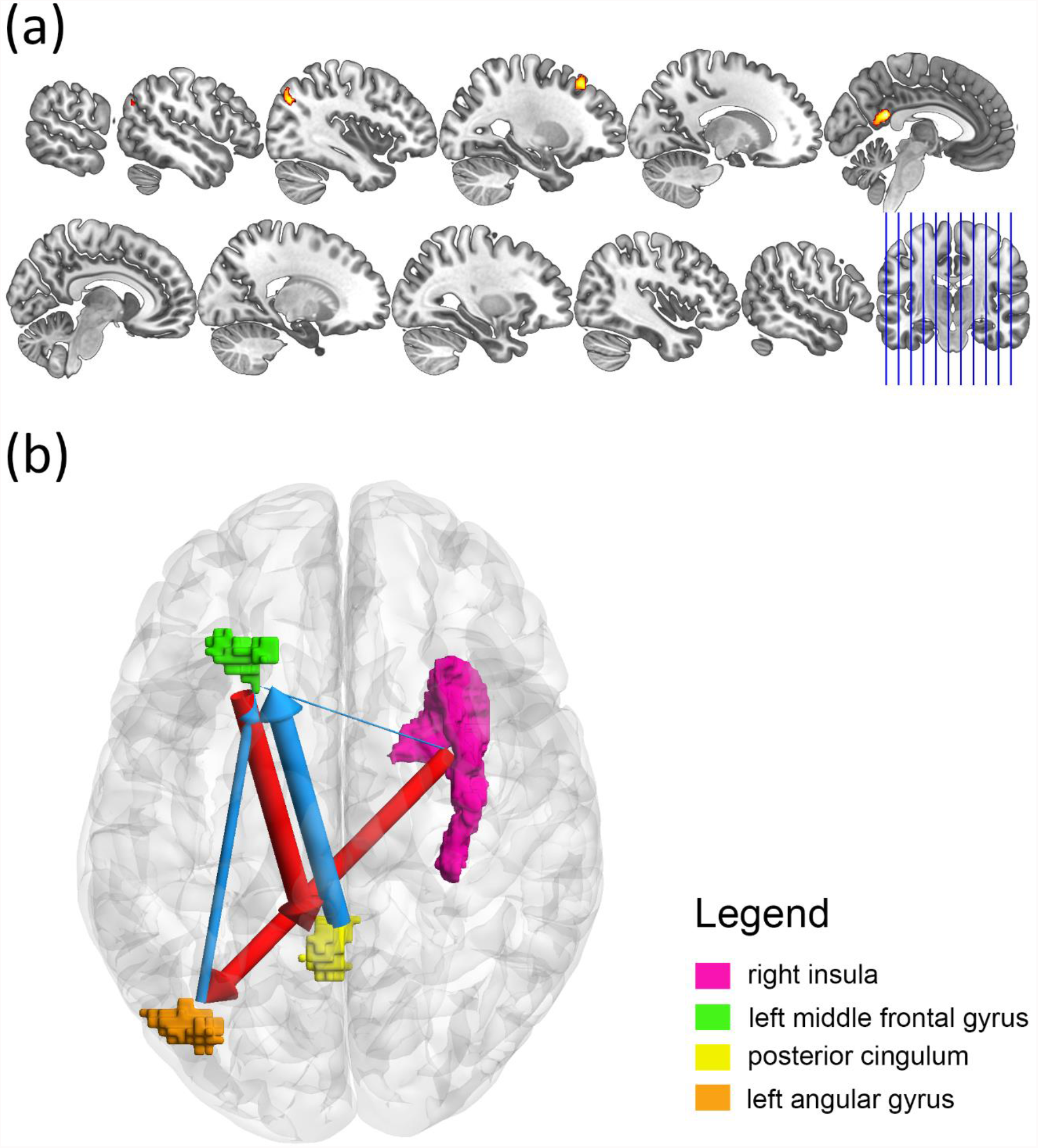
Seed-based and Effective Connectivity. (a) Regions that showed a significant group-by-time interaction with the seed region of the right insula (purple). After a minimum of 4 months an increase in connectivity was found in the posterior cingulum (p_corr_ = 0.047 | yellow), left middle frontal gyrus (p_corr_ = 0.037 |green) and left angular gyrus (p_corr_ = 0.028 | orange) for the MtF participants in comparison to the male controls. (b) Indicates the effective connectivity interaction effects in relation to the reference conditions i.e. after a minimum of 4 months of hormone therapy where MtF participants were compared to male controls. Increases in effective connectivity are indicated in red and decreases in blue. Line thickness represents the strength of the effective connectivity difference. All effects have a > 99 % posterior probability.

A minimum of 4 months GHT induced changes in the effective connectivity for the MtF participants when compared to MC. An increase in effective connectivity from the right insula to the left angular gyrus and left middle frontal gyrus to the posterior cingulum (PC) was found. Furthermore, decreases were discovered from the posterior cingulum and left angular gyrus respectively to the left middle frontal gyrus. Finally, a very slight decrease in effective connectivity from the right insula to the left middle frontal gyrus was revealed; see Fig 1b for a graphical representation.

## 4. Discussion

The current study aimed to assess changes to effective connectivity of the insula at rest under gender-affirming hormone treatment. In a seed-based analysis, we found that in comparison to cisgender males, MtF subjects showed an increase in effective connectivity from the right insula to the left AG and from the left MFG to the PC at rest after receiving GHT. In the same cohort, decreased effective connectivity could be shown both from the PC as well as the left AG to the left MFG and from the right insula to the left MFG. Here, we aim to discuss potential interpretations of our data in view of previously published findings on functional characteristics of the insula, MFG, AG and PC.

### Interoception and own-body perception

Interoception is the ability to recognize and process inner sensations of the body. It is strongly associated with own-body perception (50). As reviewed by Berntson et al., interoception is a highly complex central nervous function involving the identification, processing and integration of signals from peripheral chemo-, mechano-, osmo- and humoral receptors via different neuronal pathways, networks and circuits (51). Looking at the integration of sensory input to orchestrate behavior and attention, both the central autonomic network (see review by Beissner et al. (52) or Bonaz et al. (53)) as well as the salience network (SN) need to be taken into account (54). As suggested by Menon and Uddin, the SN segregates the most important among internal and external signals, aiming to coordinate behavior and associated reactions (55). At rest, also the default mode network (DMN) was suggested to be highly relevant for self-referential mental activity (56).

As an essential component of both the CAN (57) as well as the SN (58), the insular cortex is functionally implicated in somatosensory information processing (59) and interoception (14, 60-62). Although the SN was shown to work independently of the DMN, co-activation patterns were demonstrated between the posterior insula and the DMN (63). Here, we found an increase in effective connectivity of the insula to the left AG at rest in transgender women when compared to male controls after GHT treatment. The AG, one of the major connecting hubs within the human brain, is implicated in a large variety of processes, involving multiple aspects of higher-level cognition (64). Besides the widely known role of the angular gyrus in, e.g., autobiographic and semantic memory, arithmetic tasks, semantic processing as well as visuospatial navigation (see review by Seghier (65)), Burke and colleagues found significantly higher AG-activation in healthy adults while performing the so-called body localizer task, implemented to localize brain regions that are active while pictures of the own and other human bodies are presented and processed (66), suggesting a role of the AG in visual body perception. Own-body perception is a highly interesting task in the context of gender dysphoric patients who experience incongruence between their sex assigned at birth and their experienced gender.

Additionally, higher effective connectivity between the left MFG and the PC was shown. With connections to the CAN and as part of the salience system, the cingulum is one of the core interoceptive areas of the human brain (67). For instance, de la Fuente et al. found that grey matter volume of the left PC positively correlated with interoceptive accuracy in an addiction study where the relationship between cardiac interoception and cocaine dependence was assessed to define neurocognitive markers of interoceptive tuning using structural and functional MRI (68). Interestingly, we also found a decrease in effective connectivity, namely in the PC as well as the left AG towards the left MFG and from the right insula to the left MFG. The connectivity changes between these regions may be interpreted as a balancing communication pattern within the DMN. On the other hand, altered inter-network-communication pathways after GHT could be discussed between the DMN, CAN and SN.

The mentioned effects were detected after GHT, suggesting an influence of cross-sex hormone therapy on selected regions and networks within the brain that are centrally involved in interoceptive processing. Interoception is an aspect of paramount importance in the treatment of gender dysphoria due to its relevance for own-body perception. In a study that addressed brain network connectivity patterns involved in own-body perception across gender-dysphoric patients and cisgender controls, Feusner et al. investigated the relation of DMN and SN activity to self-perception ratings using images of the subject’s own body that were modified towards the opposite gender (69). The results pointed towards altered connectivity within networks involved in own-body perception in the context of self in GD. However, own-body perception in particular and interoception in general are not only relevant for GD, but seem to play a crucial role in a multitude of psychiatric diseases, e.g., anorexia nervosa (70), anxiety (71) and major depression (72). These conditions show higher prevalence rates in females and phenotypic differences are well documented (73) with women paying more attention to interoceptive sensations then men while showing worse interoceptive accuracy for certain tasks (74, 75).

As discussed by Murphy et al., the interaction of hormonal changes and their relation to interoception could be of interest when exploring the physiologic mechanisms behind gender differences in psychiatric conditions (76). In a study that aimed to investigate gender-related differences in functional connectivity at rest, Sie et al. revealed that females in the early-adulthood stage show stronger negative resting-state functional connectivity between the right anterior insula and the medial DMN than males, suggesting a relevance of sex when looking at resting-state network connectivity linked to interoceptive processes (77). In our sample, the long-term administration of estrogens and androgen blockers in transgender women similarly resulted in altered connectivity patterns of the insula as well as other brain regions that are implicated in DMN activity.

### Language, literacy and semantic processing

The regions exhibiting altered effective connectivity at rest after long-term feminization via estrogen treatment and androgen-blocker administration in transgender women are all known to be implicated in at least one aspect of language processing. Since gender differences in this cognitive domain were discussed in a multitude of neuroimaging and neurolinguistic publications, the effect of exogenous estrogen on language processing needs to be considered in view of the present data.

As stated by Chong and coworkers, the functional heterogeneity of the insular cortex and its activation patterns both in task-based approaches as well as resting-state analyses point towards a role as an interface coordinating the interpretation of and reaction to external stimuli involved in general cognition as well as affective and sensory-motor processing (78), which is relevant for, e.g., emotion regulation (79), goal-directed cognition (80) and linguistic processing (81). In the context of language, Ardila et al. found that both the right and the left insula were implicated in certain language-related activation patterns, such as language production, understanding and repetition as well as lexico-semantic associations (82). As reported in two meta-analyses (83, 84), also the left AG has been associated with semantic processing. Furthermore, it plays a critical role in reading comprehension (85), semantic memory (86) and understanding of speech (87). Furthermore, the left middle frontal gyrus was associated with the development of literacy (88), a region that showed an increased effective connectivity towards the posterior cingulum. The alterations in effective connectivity comprising the insula, the left AG, the left MFG, and the PC could be a direct effect of the administration of estrogen in the scope of GHT.

Despite conflicting results on gender aspects of language and the assumption that linguistic processing follows a sexually dimorphic activation pattern within the brain, many authors suggested a female advantage in language-related tasks, especially in verbal fluency tasks (89). One explanation that gained attention in this regard is the assumption that women show a more pronounced bihemispheric representation of language than men (90-92). In addition to studies on language lateralization, also gender-specific differences in intrahemispheric activation patterns were discussed (93).

Here, we suggest that the long-term administration of estrogen resulted in an increase of effective connectivity at rest within certain networks known to be involved in verbal memory and linguistic processing. These findings are in line with results from an fMRI study investigating cerebral activation in language processing, where Sommer et al. found that total language activity correlated with estrogen levels after GHT (94). Another explanation that needs to be addressed is the possible effect of androgen blockade. As reported by Cherrier et al., androgen suppression was associated with better performance in verbal memory in men with prostate cancer (95). Studies on linguistic capacities in the course of the menstrual cycle also support the idea of better language performance when estrogen levels are high, whereas androgen levels are low (96).

Since the abovementioned connectivity changes over time were only true for transgender, but not for cisgender women, it could be hypothesized that exogenously administered estrogen or androgen blockers enhanced synaptic plasticity in a multimodal manner. However, further studies need to be done to explore the underlying mechanisms, e.g., whether estrogen administration affects endothelial receptor density (97, 98), grey matter volume (99), white matter microstructure (100), alterations of intra-cerebral estrogen synthesis (101) and the interaction with certain neurotransmitter systems such as the serotonergic system (102, 103).

### Limitations

The results presented here should be seen in light of some limitations. Due to recruitment difficulties, transgender subgroups were considerably imbalanced.

Additionally, it is hardly possible to determine if the interaction effects presented here could have been influenced by mood changes and improved own-body perception in the course of transition.

## 5. Conclusion

The present study revealed that after long-term estrogen administration and anti-androgen treatment in the context of gender-affirming hormone therapy, transgender women showed significant changes of resting-state connectivity between the right insula, left AG, left MFG and PC when compared to cisgender male controls, suggesting a considerable influence of GHT on brain networks implicated in interoception, own-body perception and higher-level cognition as it is needed for language processing. Investigations considering the underlying mechanisms on a molecular level are needed to sufficiently understand the interplay of GHT, gender dysphoria and functional connectivity.

## Data Availability

Due to data protection laws, processed data is available from the authors upon reasonable request.

## 6. Acknowledgements and funding

This research was funded in whole, or in part, by the Austrian Science Fund (FWF) grant number KLI 504 to Rupert Lanzenberger., the Medical Imaging Cluster of the Medical University of Vienna, and by the grant „Interdisciplinary translational brain research cluster (ITHC) with highfield MR” from the Federal Ministry of Science, Research and Economy (BMWFW), Austria. For the purpose of open access, the author has applied a CC BY public copyright licence to any Author Accepted Manuscript version arising from this submission. MB. Reed is a recipient of a DOC fellowship of the Austrian Academy of Sciences at the Department of Psychiatry and Psychotherapy, Medical University of Vienna. We thank the graduated team members and the diploma students of the Neuroimaging Lab (NIL, headed by R. Lanzenberger) as well as the clinical colleagues from the Department of Psychiatry and Psychotherapy of the Medical University of Vienna for clinical and/or administrative support. In particular, we would like to thank B. Spurny-Dworak as well as R. Seiger for technical support and V. Ritter for administrative help.

